# Sex differences in the pleiotropy of hearing difficulty with imaging-derived phenotypes: a brain-wide investigation

**DOI:** 10.1101/2023.08.25.23294639

**Authors:** Jun He, Brenda Cabrera-Mendoza, Flavio De Angelis, Gita A. Pathak, Dora Koller, Sharon G. Curhan, Gary C. Curhan, Adam P. Mecca, Christopher H. van Dyck, Renato Polimanti

## Abstract

**Background:** Hearing difficulty (HD) is one of the major health burdens in older adults. While aging-related changes in the peripheral auditory system play an important role, genetic variation associated with brain structure and function could also be involved in HD predisposition.

**Methods:** We analyzed a large-scale HD genome-wide association study (GWAS; N_total_ = 501,825, 56% females) and GWAS data related to 3,935 brain imaging-derived phenotypes (IDPs) assessed in up to 33,224 individuals (52% females) using multiple magnetic resonance imaging (MRI) modalities. To investigate HD pleiotropy with brain structure and function, we conducted genetic correlation, latent causal variable (LCV), Mendelian randomization (MR), and multivariable generalized linear regression analyses. Additionally, we performed local genetic correlation and multi-trait colocalization analyses to identify genomic regions and loci implicated in the pleiotropic mechanisms shared between HD and brain IDPs.

**Results:** We observed a widespread genetic correlation of HD with 120 IDPs in females, 89 IDPs in males, and 171 IDPs in the sex-combined analysis. The LCV analyses showed that some of these genetic correlations could be due to cause-effect relationships. For seven correlations, the causal effects were also confirmed by the MR approach: vessel volume→HD in the sex-combined analysis; hippocampus volume→HD, cerebellum grey matter volume→HD, primary visual cortex volume→HD, and HD→rfMRI-ICA100 node 46 in females; global mean thickness→HD and HD→mean orientation dispersion index in superior corona radiata in males. The local genetic correlation analyses identified 13 pleiotropic regions between HD and these seven IDPs. We also observed a colocalization signal for the rs13026575 variant between HD, primary visual cortex volume, and *SPTBN1* transcriptomic regulation in females.

**Conclusion:** Brain structure and function may have a role in the sex differences in HD predisposition via possible cause-effect relationships and shared regulatory mechanisms.

## BACKGROUND

Hearing difficulty (HD) is a common health condition. According to estimates, 1.57 billion people had HD in 2019, causing a global cost of about one trillion dollars due to losses of quality of life and productivity^1–3^; the affected population was predicted to be 2.45 billion by 2050^2^. Due to the rapid aging of the world’s population and increased life expectancy, the years lived with disability (YLDs) attributed to HD increased by 73.6% from 1990 to 2019. HD is currently the third leading cause of YLDs for all ages and the largest cause for people aged 70 and over^2,4^. HD can affect language abilities, psychosocial health, quality of life, educational level, and economic independence^5,6^ and is associated with adverse health consequences, including neurodegenerative, respiratory, psychiatric, and cardiometabolic diseases^7–9^.

Our previous study highlighted the association between the genetic basis of HD and brain transcriptomic regulation^10^. Based on this, we hypothesized that the brain structure and function have a role in the development of HD. The risk factors of HD, including genetic predisposition, certain health conditions, adverse lifestyle behaviors, and environmental exposures experienced across the lifespan, mainly affect the functions of the peripheral auditory system, cochlear nerves, and brainstem auditory pathways^11^. In addition, central nervous system components involved in information processing, cognition, and other processes may also be involved^8^. Several studies have identified the relationships between hearing-related traits and brain structure, including grey matter volume, white matter volume, and cortical thickness across temporal, limbic, frontal, and occipital areas^12–17^. Moreover, fractional anisotropy of the auditory areas and the splenium of the corpus callosum and activation of auditory areas, visual association areas, and attention networks were reported to be associated with HD^18,19^. However, those studies have mainly analyzed the association of HD with different brain magnetic resonance imaging (MRI) traits or only the correlation between them, lacking evidence of the brain structure and function contributing to HD development. Most of these studies investigated a limited number of brain MRI or functional MRI phenotypes without a thorough analysis across the whole brain structure and function. Since only phenotypic data and regression methods were used when assessing the associations, to our knowledge no studies leveraged genome-wide data and analytical approaches to elucidate these relationships and the possible underlying mechanisms. This is an important gap, because genetically informed causal inference analyses are less susceptible to potential confounding and reverse causation bias.

Large-scale biomedical databases like the UK Biobank (UKB) have recruited substantial numbers of participants and provide genome-wide information. These genetic data coupled with newly developed analytical approaches permit us to evaluate causal relationships between complex traits that are difficult to assess using traditional randomized controlled trial designs. In the current study, we used genome-wide association data related to 3,935 brain imaging-derived phenotypes (IDPs) and HD from several large datasets to investigate HD pleiotropy with brain structure and function and their causal relationships as well as to identify genomic regions implicated in their shared pleiotropic mechanisms.

## METHODS

### Data sources

We leveraged a genome-wide association study (GWAS) meta-analysis of HD and GWAS of 3,935 IDPs to explore both sex-combined and sex-stratified pleiotropy between HD and IDPs. In brief, the meta-analysis for HD pooled data from the UKB^20^, Nurses’ Health Study (NHS) I^21^, NHS II^21^, and Health Professionals Follow-Up Study (HPFS)^22^, which included 501,825 unrelated adults - 56% of them were females^10^. The UKB is a large population-based prospective cohort including deep genetic information and health conditions data from over 500,000 UK participants (40–69 years old, 54% were females). In the UKB, HD was assessed by a touchscreen question “Do you have any difficulty with your hearing?” (UKB Field ID: 2247). The most recent assessment was used in the GWAS when multiple assessments were available and those who were completely deaf were excluded^10^. The other three cohorts enrolled 121,700 (NHS I: 30–55 years old) and 116,430 (NHS II: 25–42 years old) female registered nurses and 51,529 male health professionals (HPFS: 40–74 years old). In each study, a questionnaire was used to collect the hearing status of participants. Cases were defined when mild, moderate, severe (non-hearing aid user), or severe (hearing aid user) HD were reported^10^. Detailed information about the meta-analyzed data of HD can be found elsewhere^10^. In this study, genome-wide association statistics related to 3,935 IDPs were assessed in up to 33,224 individuals (52% females) by the UKB team. A full description of each brain IDP is available in Supplementary Table S1. Brain MRI was performed using Siemens Skyra 3 T scanners. Brain structural, diffusion, and functional phenotypes derived from T1-weighted structural image, T2-weighted fluid-attenuated inversion recovery (FLAIR) structural image, diffusion MRI (dMRI), resting-state functional MRI (rfMRI), task functional MRI (tfMRI), and susceptibility-weighted imaging (SWI) modalities were classified into 17 categories. Detailed imaging processing, genetic pre-processing, and quality control information were described previously (https://biobank.ctsu.ox.ac.uk/crystal/crystal/docs/brain_mri.pdf)^23^.

### Linkage disequilibrium score regression

We used linkage disequilibrium score regression (LDSC) to calculate SNP-based heritability of each IDP and genetic correlations (r_g_) between HD and 3,935 IDPs for both sex-combined and sex-stratified analyses^24,25^. The genetic correlation estimates are not biased by sample overlap when using LDSC approach^25^. Pre-computed linkage disequilibrium (LD) scores estimated using the 1000 Genomes European data were used in the regression^26^. LDSC was performed only based on SNPs from Hapmap3^27^, INFO > 0.9, and minor allele frequency (MAF) > 0.01 according to the approach default settings. Due to the modest sample size of IDP GWAS (up to 33,224) and the relative lack of genome-wide significant SNPs (*P* < 5 × 10^-8^) for specific IDPs, we used a nominally significant threshold (*P* < 0.05) to identify IDPs genetically correlated with HD.

### Latent causal variable analysis

To assess whether the genetic correlations between IDPs and HD defined by LDSC were due to possible causal relationships, we performed the latent causal variable (LCV) analysis to estimate the genetic causality proportions (gcp) between the two phenotypes^28^. LCV considers the genetic correlation between two traits (trait 1 and trait 2) to be mediated by a latent variable that has a causal effect on each trait. This method uses cross-trait genetic correlations estimated from LDSC and quantifies the causal relationship by the gcp values. The gcp ranges from −1 to 1 where 0 indicates no genetic causality, −1 or 1 indicates full genetic causality, and other values indicate partial genetic causality between the two traits. The sign of gcp is related to the causal direction between the traits, with positive values indicating that trait 1 is the causal factor leading to trait 2 as the outcome, while negative gcp values correspond to a causal effect from trait 2 to trait 1. LCV also calculates the genetic correlations between the two traits by LDSC. We used pre-computed LD scores computed using the 1000 Genomes phase 3 European data when evaluating gcp estimates. The statistically significant threshold of gcp was defined using Bonferroni multiple testing correction accounting for the number of traits analyzed.

### Mendelian randomization

MR was used to verify the causal relationships identified by LCV analysis and estimate the effects both from IDPs to HD and from HD to IDPs. Because of the existing sample overlap between IDPs and HD GWAS due to the presence of UKB participants in both analyses, we performed an inverse variance weighting (IVW) MR analysis using the MRlap approach^29^. This is an extension of the two-sample MR method^30^ that provides a corrected causal effect estimate for the relationship of two traits, by accounting for potential sample overlap, as well as weak instrument bias and winner’s curse simultaneously^31^. A *P*-value threshold of 1 × 10^-5^ was chosen to select instrumental variables (IVs) due to the modest sample size and the relative lack of genome-wide significant associations with brain IDPs. The default setting of distance pruning (500 kb) in MRlap package was used to identify independent IVs, which is equal to the LD pruning method (LD cutoff = 0)^29^. Additionally, to avoid the reverse causal association (the outcome causes the exposure), MRlap only included SNPs that were more strongly associated with the exposure compared with the outcome as determined by Steiger’s test (*P* < 0.05)^32^. The 1000G LD scores were included in the LDSC analyses. As recommended for MR analyses^33^, we avoided inference based simply on *P*-value thresholds. The direction and strength of the effects estimated via analyses, together with the corresponding *P*-values, were considered to better reflect the spectrum of evidence related to the MR results.

### Multivariable generalized linear regression

In the generalized linear regression analysis, we selected unrelated participants with European ancestry in UKB to estimate the associations between IDPs and HD. We developed three models for each causal relationship filtered out in the MR analysis. In model 1, no covariate was considered. In model 2, we adjusted for age, sex, and age × sex. In model 3, age^2^ and age^2^ × sex were included. In the female- and male-specific analyses, covariates related to sex were not included. We compared the size and sign of the effect estimates between generalized linear regression and MR analysis. In all three models, IDP information was extracted from the imaging visit, and HD data was obtained from self-reported hearing difficulty (UKB Field ID: 2247). The individuals having missing values for HD, IDP, age, and sex were excluded from the analysis. Logistic regression was used when testing the association of IDPs with HD, while linear regression was chosen when testing that of HD with IDPs. The analysis was performed using glm function in R and associations with *P* < 0.05 were considered statistically significant.

### Local analysis of [co]variant association

Local analysis of [co]variant association (LAVA) was used to estimate local genetic correlations between IDPs and HD^34^. The analysis was conducted across 2,495 semi-independent genetic loci (∼1Mb for each locus) partitioning the autosomal genome while minimizing LD between the loci. To ensure stable and interpretable local genetic correlations with sufficient genetic signals, we performed a two-step analytical strategy for LAVA. In the first step, we used univariate analysis as a filtering method to select loci with significant local heritability (*P* < 0.05) for seven IDPs identified in the MR analysis. In the second step, we performed bivariate analysis across all genetic loci selected in the first step to identify significant local genetic correlation between IDPs and HD using false discovery rate (FDR) to correct the threshold for multiple testing (FDR *q* < 0.05). Sample overlap was adjusted using the intercepts of bivariate LDSC, and the European panel of phase 3 of 1000 Genomes (MAF > 0.5%) was used as LD reference panel.

### Multi-trait colocalization analysis

To elucidate the colocalization among IDPs, HD, and brain transcriptomic regulation, we used a multi-trait colocalization approach (HyPrColoc)^35^ assessing the shared genetic etiology and prioritizing causal variants. The analysis was conducted within the same 2,495 loci defined in LAVA and across the seven associations identified in the MR analysis. HyPrColoc mainly provides three values, including the posterior probability of full colocalization (PPFC, i.e., a probability that all phenotypes share a causal genetic variant in a specific region), the regional association probability (P_R_, i.e., a probability indicates all phenotypes share one or more variants in a specific region), and the proportion the PPFC explained by the candidate causal variant (P_snp_). After identifying colocalized traits and their subsequent shared causal variants, we also extracted their expression quantitative trait loci (eQTL) statistics from the GTEx v.8 release^36^ (https://gtexportal.org/home/) to test their colocalization with brain-specific transcriptomic gene regulation. We only selected genes associated with the causal variants of IDPs and HD (m-value > 0.9) in brain tissues to conduct this analysis^37^. In all analyses conducted, the default parameters of HyPrColoc package were used and the sample overlap between traits was adjusted using data of pair-wise marginal correlations between the traits, LD matrix in the region, and proportion of sample overlap.

## RESULTS

### Global genetic correlation

LDSC analysis showed that the global genetic correlation between HD and 3,935 IDPs ranged from −0.58 to 0.49 in the sex-combined model, −0.78 to 0.70 in females, and −0.67 to 0.68 in males (Fig. 1; Supplementary Tables S2-S4). Although the genetic correlation estimates were mostly consistent between females and males (r_Pearson_ = 0.21, *P* < 2.2 × 10^-16^), we found three IDPs showing significant sex differences after Bonferroni correction (*P* < 0.05/3,024; Supplementary Fig. S1; Supplementary Table S5). They were all related to rfMRI connectivity: ICA100 edge 714 (IDP 3142; female r_g_=-0.78, male r_g_=0.06, sex-difference *P* = 1.69 × 10^-7^), ICA100 edge 955 (IDP 3383; female r_g_=-0.53, male r_g_=0.06, sex-difference *P* = 1.88 × 10^-6^), and ICA100 edge 1371 (IDP 3799; female r_g_=-0.50, male r_g_=0.04, sex-difference *P* = 1.48 × 10^-5^). Considering IDPs with SNP-based heritability Z > 4, we found 171, 120, and 89 IDPs having nominally significant genetic correlations (*P* < 0.05) with HD for the sex-combined, female, and male analyses, respectively. In the sex-combined analysis, the top significant genetic correlation was for BA-exvivo rh area BA2 (IDP 0797; r_g_ = −0.17, *P* = 4 × 10^-4^), followed by IDP dMRI TBSS OD Retrolenticular part of internal capsule R (IDP 1997; r_g_ = −0.13, *P* = 5.00 × 10^-4^), aparc-a2009s rh thickness S-cingul-Marginalis (IDP 1297; r_g_ = 0.17, *P* = 8.00 × 10^-4^), rfMRI connectivity ICA100 edge 604 (IDP 3032; r_g_ = −0.19, *P* = 1.40 × 10^-3^), and aparc-a2009s rh volume S-parieto-occipital (IDP 0638; r_g_ = 0.12, *P* = 2.10 × 10^-3^). In females, four of the top five IDPs genetically correlated with HD were related to cortical thickness, while in males, all of the top five were dMRI white matter tract IDPs (Fig. 1; Supplementary Tables S2-S4).

**Fig. 1.**
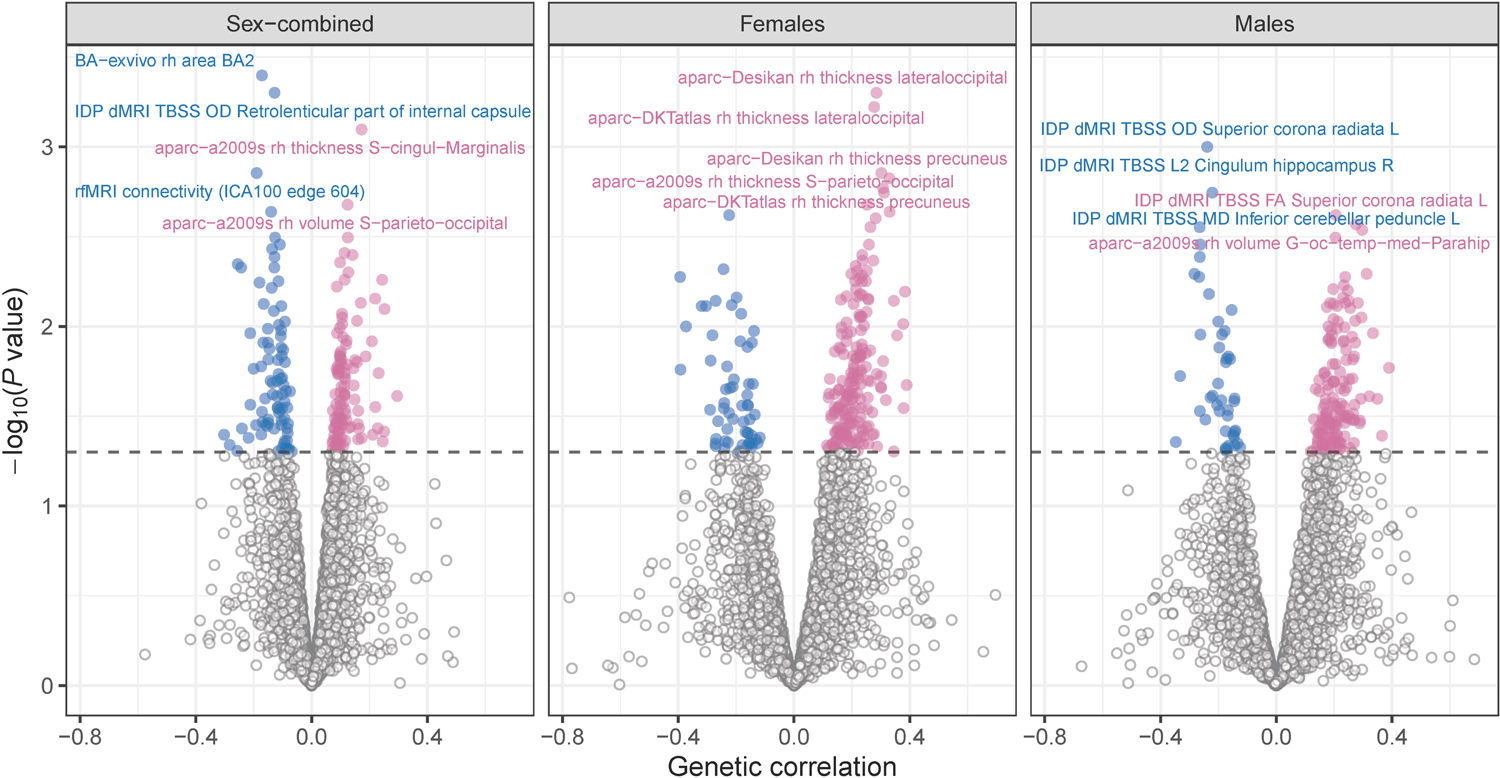
Global genetic correlation between hearing difficulty (HD) and brain imaging-derived phenotypes (IDPs) in the sex-combined and sex-stratified analyses. The dashed line refers to the nominal significant threshold with *P* = 0.05. Blue points indicate the IDPs having a significant negative genetic correlation with HD, while red points indicate those having a significant positive genetic correlation with HD. Labels are reported for the top five IDPs with the strongest HD genetic correlation. Full results are available in Supplementary Tables S2-S4.

### Latent causal variable analysis

To assess possible causal effects underlying the genetic correlations using the LDSC model, we performed the LCV analysis and calculated gcp for the identified associations. In this analysis, IDPs were set as trait 1 and HD as trait 2, whereby a positive gcp indicated a causal effect from IDP to HD. Conversely, negative gcp values corresponded to a causal effect from HD to IDP. After Bonferroni correction (*P* < 0.05/171), five potential causal relationships between IDPs and HD were observed in the sex-combined analysis (Fig. 2, Supplementary Table S6): IDP T1 FAST ROIs R supramarg gyrus post (IDP 0065; gcp = 0.807, *P* = 2.29 × 10^-12^), aparc-a2009s rh volume G-temp-sup-Plan-polar (IDP 0608; gcp = 0.341, *P* = 5.14 × 10^-11^), rfMRI amplitudes ICA100 node 27 (IDP 2190; gcp = 0.318, *P* = 1.32 × 10^-10^), rfMRI connectivity ICA100 edge 1140 (IDP 3568; gcp = 0.633, *P* = 7.71 × 10^-9^), and aseg rh volume vessel (IDP 0220; gcp = 0.460, *P* = 2.88 × 10^-7^). Twenty-seven significant potential causal relationships (*P* < 0.05/120) were suggested in the female-specific analysis, with the top significance being IDP dMRI TBSS MO Medial lemniscus R (IDP 1535; gcp = −0.133, *P* = 6.44 × 10^-40^), followed by three regional and tissue volume IDPs (Fig. 2, Supplementary Table S7). Fifteen potential causal relationships (*P* < 0.05/89) were suggested in the male-specific analysis (Fig. 2, Supplementary Table S8). The top five IDPs were both white matter tract IDPs with three of them being related to cingulum hippocampus. Among all 47 significant IDPs identified by LCV analysis, 41 had potential causal effects on HD, with the remaining six IDPs being possible outcomes of HD (Fig. 2).

**Fig. 2.**
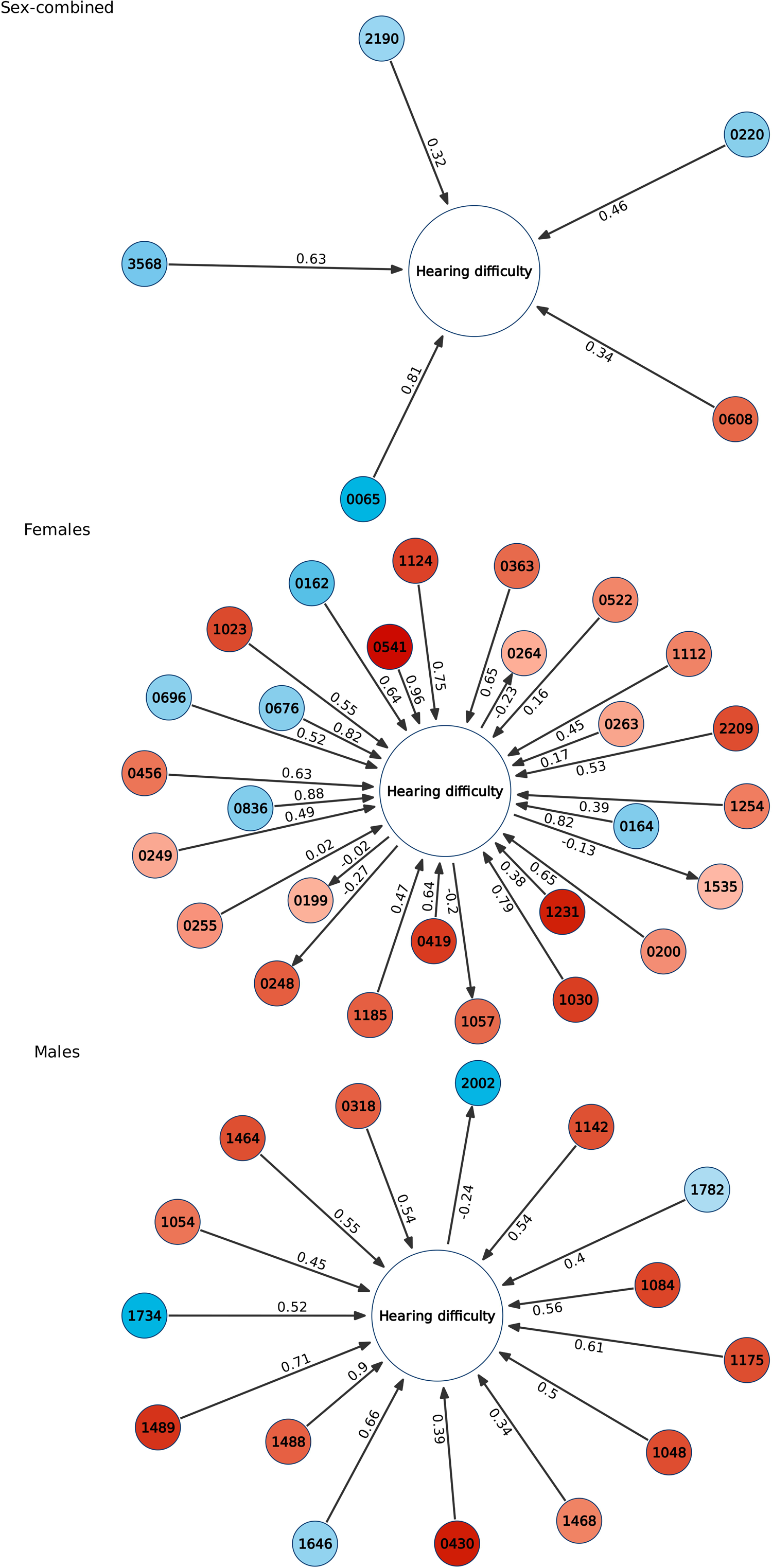
Putative causal effects between hearing difficulty (HD) and brain imaging-derived phenotypes (IDPs) identified by latent causal variable model after Bonferroni multiple testing correction. The arrows indicate directions of the causal effect with the numbers alongside it being the genetic causality proportions. The numbers in the circles respond to IDP IDs. The colors filled in the circles map the genetic correlation estimates between HD and IDPs, with blue representing negative correlation and red representing positive correlation (darker color representing stronger correlation). 0608: aparc-a2009s rh volume G-temp-sup-Plan-polar; 0220: aseg rh volume vessel; 2190: rfMRI amplitudes (ICA100 node 27); 3568: rfMRI connectivity (ICA100 edge 1140); 0065: IDP T1 FAST ROIs R supramarg gyrus post; 0162: IDP T1 FAST ROIs L cerebellum X; 0164: IDP T1 FAST ROIs R cerebellum X; 0199: aseg lh volume Hippocampus; 0200: aseg lh volume Amygdala; 0248: HippSubfield lh volume presubiculum-head; 0249: HippSubfield lh volume CA1-head; 0255: HippSubfield lh volume CA3-body; 0263: HippSubfield lh volume Whole-hippocampal-head; 0264: HippSubfield lh volume Whole-hippocampus; 0363: aparc-Desikan lh volume pericalcarine; 0419: BA-exvivo lh volume V1; 0456: aparc-DKTatlas lh volume pericalcarine; 0522: aparc-a2009s lh volume G-oc-temp-med-Parahip; 0541: aparc-a2009s lh volume Pole-occipital; 0676: aparc-Desikan lh area superiorparietal; 0696: aparc-Desikan rh area middletemporal; 0836: aparc-DKTatlas lh area superiorparietal; 1023: aparc-Desikan lh thickness caudalmiddlefrontal; 1030: aparc-Desikan lh thickness lateraloccipital; 1057: aparc-Desikan rh thickness caudalmiddlefrontal; 1112: BA-exvivo rh thickness V2; 1124: aparc-DKTatlas lh thickness lateraloccipital; 1185: aparc-a2009s lh thickness G+S-cingul-Mid-Post; 1231: aparc-a2009s lh thickness S-front-sup; 1254: aparc-a2009s rh thickness G+S-paracentral; 1535: IDP dMRI TBSS MO Medial lemniscus R; 2209: rfMRI amplitudes (ICA100 node 46); 0318: ThalamNuclei rh volume VLa; 0430: BA-exvivo rh volume BA6; 1048: aparc-Desikan lh thickness superiorparietal; 1054: aparc-Desikan rh thickness GlobalMeanThickness; 1084: aparc-Desikan rh thickness supramarginal; 1142: aparc-DKTatlas lh thickness superiorparietal; 1175: aparc-DKTatlas rh thickness supramarginal; 1464: IDP dMRI TBSS FA Superior cerebellar peduncle R; 1468: IDP dMRI TBSS FA Anterior limb of internal capsule R; 1488: IDP dMRI TBSS FA Cingulum hippocampus R; 1489: IDP dMRI TBSS FA Cingulum hippocampus L; 1646: IDP dMRI TBSS MD Uncinate fasciculus R; 1734: IDP dMRI TBSS L2 Cingulum hippocampus R; 1782: IDP dMRI TBSS L3 Cingulum hippocampus R; 2002: IDP dMRI TBSS OD Superior corona radiata L. IDP description is available in Supplementary Table S1 and full results of latent causal variable model are available in Supplementary Tables S6-S8.

### Mendelian randomization and multivariable generalized linear regression analyses

We performed MR analysis to verify the causal relationships and estimate the causal effect sizes identified by the LCV model. Both directions from IDPs to HD and from HD to IDPs were assessed. In the sex-combined analysis, MR analysis confirmed the causal relationship for aseg rh volume vessel (Direction: IDP 0220→HD, Effect = −0.191 ± 0.078 (estimate ± standard error)) (Fig. 3, Supplementary Table S9). In females, four IDPs having possible causal relationships with HD were verified (Fig. 3, Supplementary Table S10), including aseg lh volume Hippocampus (IDP 0199→HD, Effect = −0.020 ± 0.008), IDP T1 FAST ROIs R cerebellum X (IDP 0164→HD, Effect = −0.035 ± 0.015), BA-exvivo lh volume V1 (IDP 0419→HD, Effect = 0.030 ± 0.014), and rfMRI amplitudes ICA100 node 46 (HD→IDP 2209, Effect = 0.239 ± 0.116). In males, MR analyses confirmed the causal effect of two IDPs (Fig. 3, Supplementary Table S11): aparc-Desikan rh thickness GlobalMeanThickness (IDP 1054→HD, Effect = 0.032 ± 0.014), and IDP dMRI TBSS OD Superior corona radiata L (HD→IDP 2002, Effect = −0.286 ± 0.134).

**Fig. 3.**
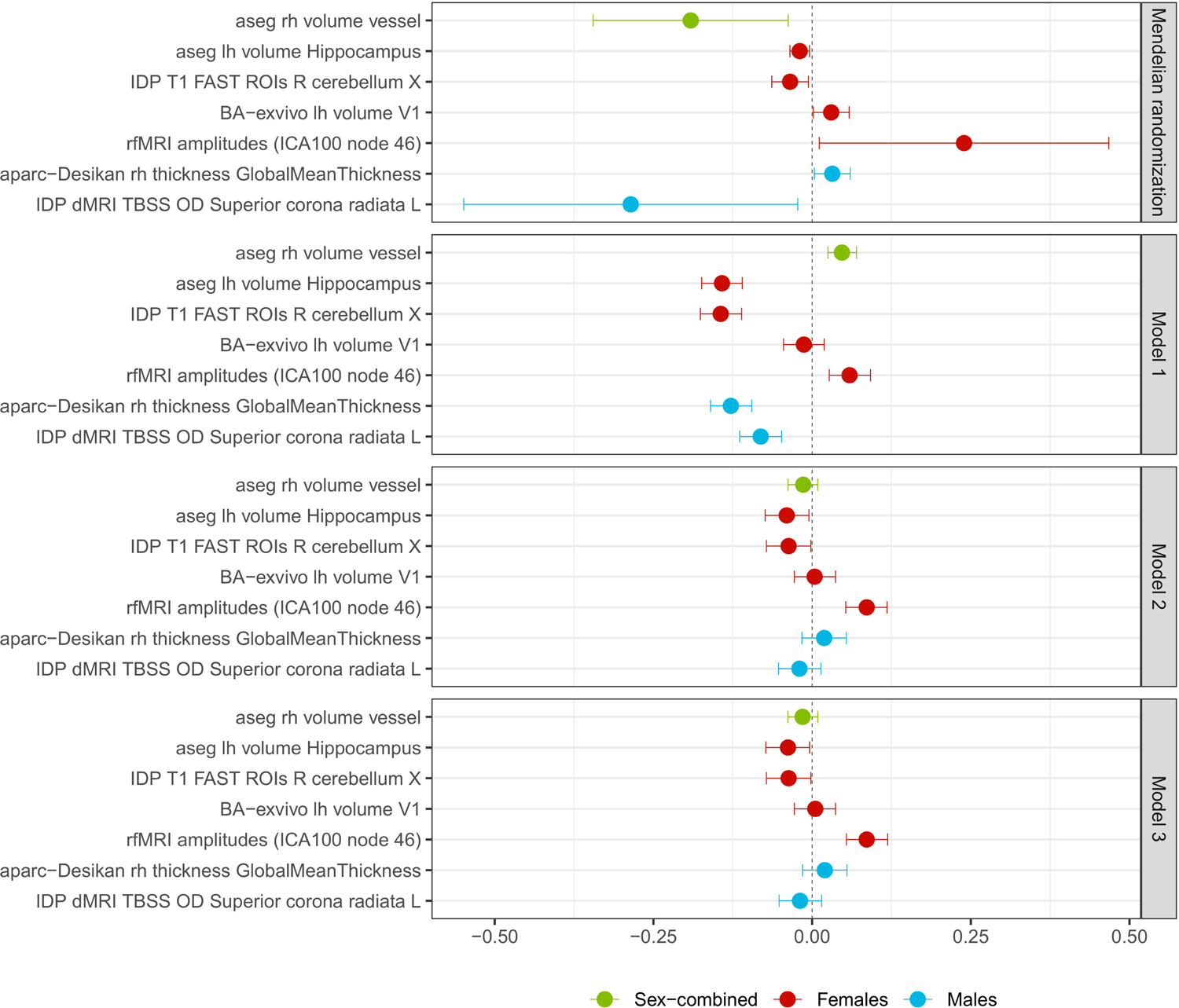
Genetically-inferred effects between hearing difficulty (HD) and brain imaging-derived phenotypes (IDPs) (Mendelian randomization *P* < 0.05) compared to phenotypic associations estimated using the UKB data. The green, red, and blue colors refer to the beta and 95%CI for the sex-combined, female, and male analyses, respectively. In generalized linear regression model 1, no covariate is included. In model 2, age, sex, and age × sex are adjusted for (only age for sex-stratified analysis). In model 3, age^2^ and age^2^ × sex were added together with the model-2 covariates (only age and age^2^ for sex-stratified analysis). Full results are available in Supplementary Tables S9-S12.

Using individual-level IDP and HD data available from the UKB, we performed generalized linear regression models to investigate further the seven IDPs identified as potentially causally linked to HD by both MR and LCV analyses. In model 1 (no covariates included), six IDPs confirmed their associations with HD (Fig. 3, Supplementary Table S12). In model 2 and model 3, the association of aseg lh volume Hippocampus (IDP 0199), IDP T1 FAST ROIs R cerebellum X (IDP 0164), and rfMRI amplitudes ICA100 node 46 (IDP 2209) with HD in females remained significant (*P* < 0.05). Overall, after accounting for covariates related to sex and age (i.e., models 2 and 3), the direction of the phenotype-level analysis was consistent with the genetically inferred effects obtained from the MR analysis.

### Local genetic correlation

Applying the LAVA approach, we estimated the local genetic correlations between HD and the seven IDPs identified in MR. The analysis was limited to genomic regions with evidence of local heritability for both HD and brain IDPs (sex-combined N = 448; female N = 2,739; male N = 1,248). In the bivariate analysis, we identified 13 genomic regions with statistically significant evidence of local genetic correlation between HD and IDPs (FDR *q* < 0.05; Fig. 4, Supplementary Table S13). In the sex-combined analysis, we observed significant local genetic correlation of HD with aseg rh volume vessel (IDP 0220) at locus 965 (chr6: 32,586,785–32,629,239; local r_g_ = 0.942, FDR *q* = 8.796 × 10^-11^), locus 966 (Chr6: 32,629,240–32,682,213; local r_g_ = 0.881, FDR *q* = 0.010), and locus 1280 (Chr8: 36641175–38,803,980; local r_g_ = −0.950, FDR *q* = 0.038). In females, the most significant local genetic correlation was detected for BA-exvivo lh volume V1 (IDP 0419) at locus 746 (Chr4: 163937104–164781847; local r_g_ = −0.853, FDR *q* = 0.009), followed by IDP T1 FAST ROIs R cerebellum X (IDP0164) at locus 103 (Chr 1: 117,046,312–118,118,667), aseg lh volume Hippocampus (IDP 0199) at locus 1844 (Chr12: 115,439,452–116,197,675), IDP T1 FAST ROIs R cerebellum X (IDP 0164) at locus 2071 (Chr15: 67,396,521–69,089,815), and rfMRI amplitudes ICA100 node 46 (IDP 2209) at locus 2224 (Chr17: 66,757,557–68,176,219). In males, the most significant local genetic correlation was for IDP dMRI TBSS OD Superior corona radiata L (IDP 2002) at locus 2465 (Chr22: 19,635,655–20,969,184; local r_g_ = −0.559, FDR *q* = 0.020), followed by aparc-Desikan rh thickness GlobalMeanThickness (IDP 1054) at locus 123 (Chr1: 165,553,813–166,458,031), locus 987 (Chr6: 52598880–53425492), locus 393 (Chr2: 215,899,571–217,566,011), and locus 2345 (Chr19: 39,370,420–40,171,413).

**Fig. 4.**
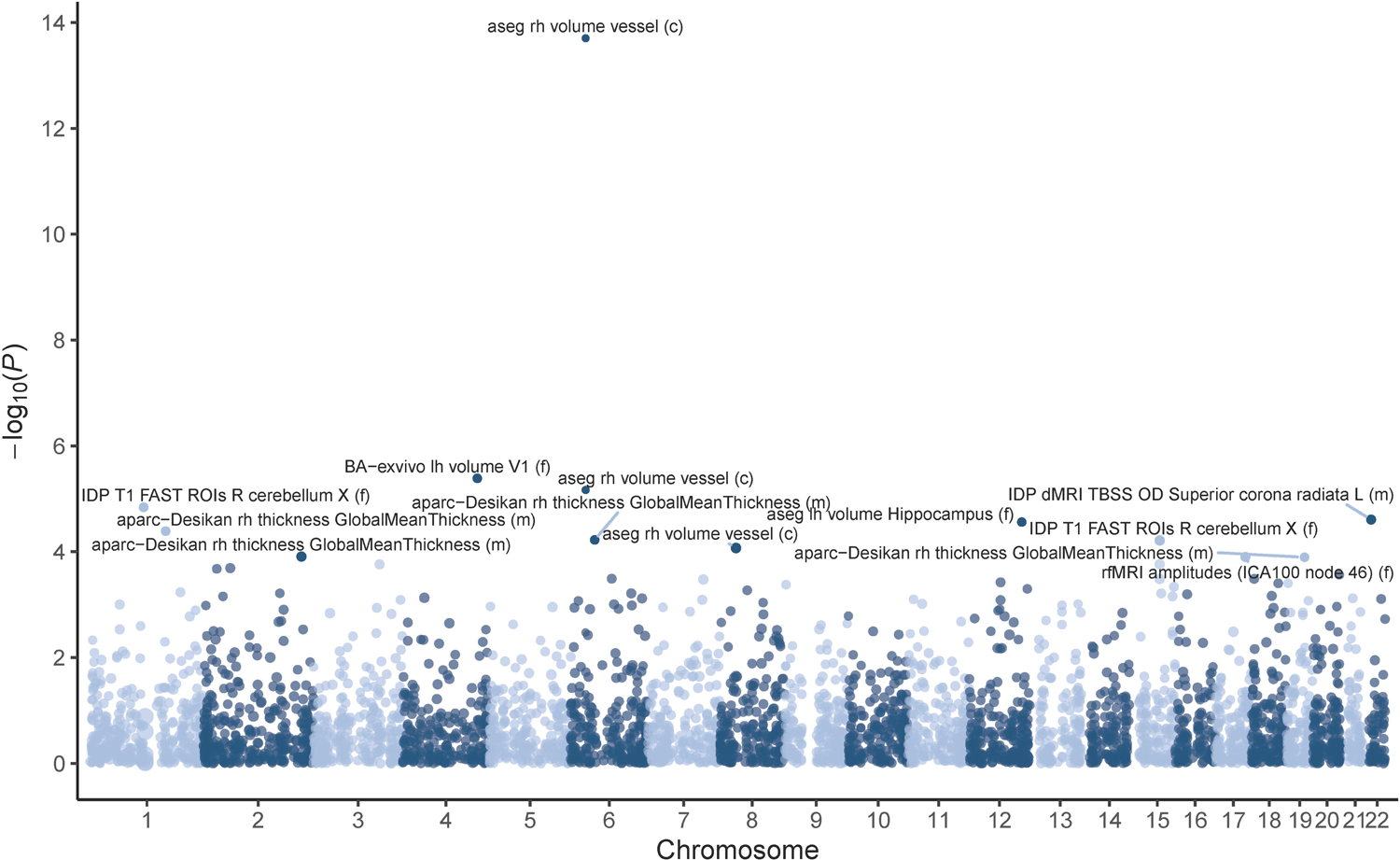
Local genetic correlation between hearing difficulty (HD) and brain imaging-derived phenotypes (IDPs) identified in the Mendelian randomization analysis. Labels are reported for HD-IDP local genetic correlation surviving false discovery rate correction (FDR *q* < 0.05). c: the sex-combined; f: females; m: males. Full results are available in Supplementary Table S13.

### Colocalization Analysis

We used HyPrColoc analysis to test the shared causal SNP between HD and the seven IDPs identified in MR analysis across the 2,495 genomic regions defined by LAVA. In females, we identified two SNPs with colocalization evidence between HD and certain brain IDPs (Supplementary Table S14): rs5899177 for aseg lh volume Hippocampus (IDP 0199; PPFC = 0.5611, P_R_ = 0.5611, P_snp_ = 0.8186) and rs13026575 for BA-exvivo lh volume V1 (IDP 0419; PPFC = 0.6598, P_R_ = 0.7655, P_snp_ = 0.9996). No significant colocalization signal was found in the sex-combined and male analyses. Adding eQTL for brain tissues with m-value > 0.9 (Supplementary Table S15) with respect to the two SNPs identified in the female-specific analysis, rs13026575 colocalized with BA-exvivo lh volume V1 (IDP 0419), and transcriptomic regulation of coding gene *SPTBN1* in three brain tissues (cerebellar hemisphere: PPFC = 0.4617, P_R_ = 0.6173, P_snp_ = 1.0000; brain cortex: PPFC = 0.5920, P_R_ = 0.7075, P_snp_ = 1.0000; nucleus accumbens basal ganglia: PPFC = 0.6613, P_R_ = 0.7723, P_snp_ = 1.0000) and non-coding gene *RPL23AP32* in brain cortex (PPFC = 0.4916, P_R_= 0.6184, P_snp_ = 1.0000) (Fig. 5, Supplementary Tables S16, S17).

**Fig. 5.**
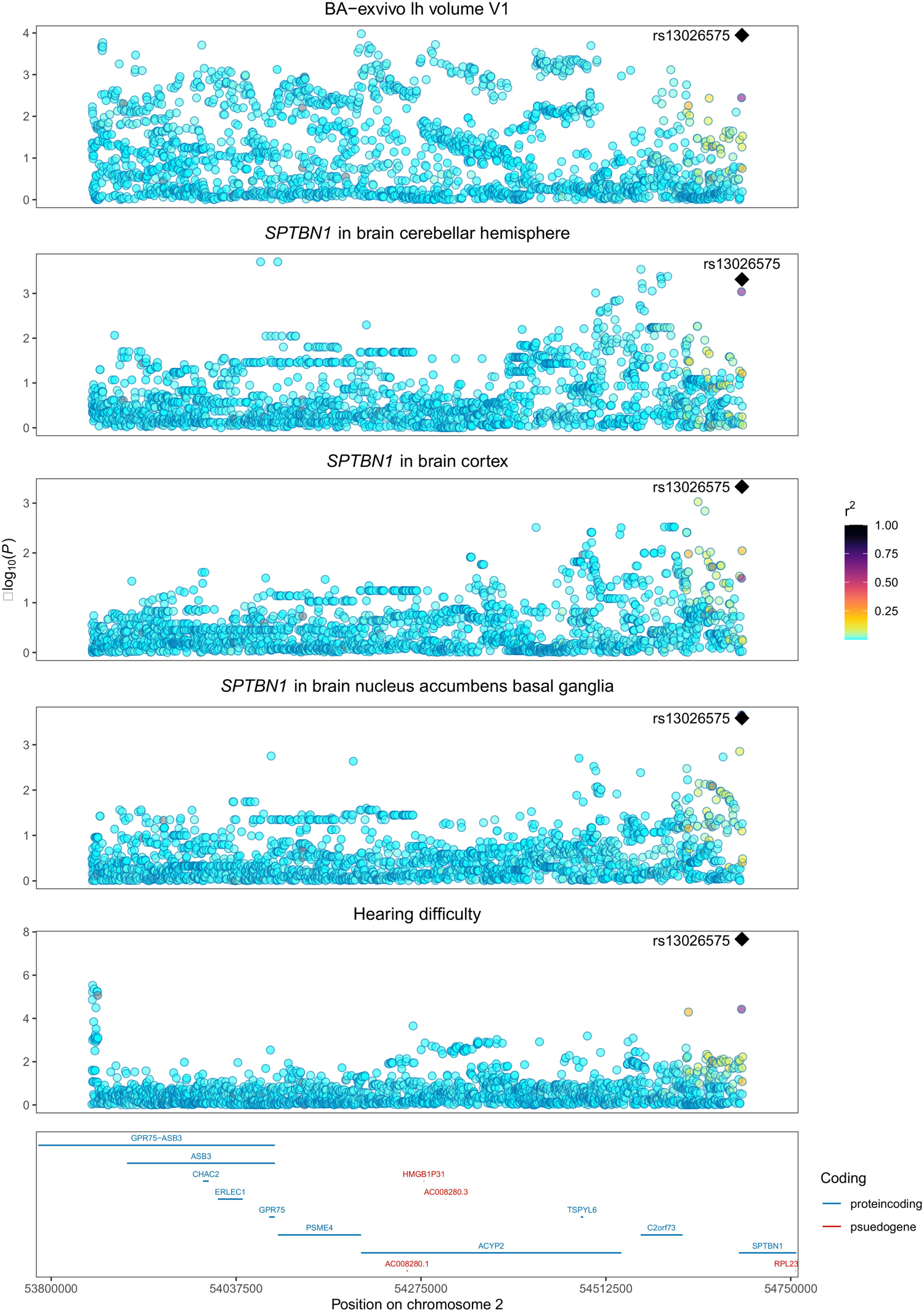
Regional association at chr2:53852121-54688234 locus. SNPs are represented by points colored relative to linkage disequilibrium (LD) r^2^. The rs13026575 is detected as a shared causal SNP for hearing difficulty (females), brain imaging-derived phenotype of volume of left primary visual cortex (IDP 0419, BA−exvivo lh volume V1, females), and *SPTBN1* transcriptomic regulation in three brain tissues from GTEx v.8 release. Full results are available in Supplementary Table S17.

## DISCUSSION

HD is a common health condition with considerable impact on psychosocial well-being, quality of life, educational attainment, and economic independence^5^. In the present study, leveraging GWAS data from four large cohorts, we provided a comprehensive assessment of the pleiotropy linking HD to changes in brain structure and function. Although we observed a certain consistency in HD-IDP genetic correlation between females and males, three rfMRI connectivity IDPs had significantly different directions of r_g_ with HD, with females more inclined to have a negative genetic correlation. These discrepancies may be related to the sex differences observed in human brain connectivity. A study of sex differences in the structural connectome of the human brain demonstrated that males had greater within-hemispheric connectivity, modularity, and transitivity in supratentorial regions, while females had predominantly between-hemispheric connectivity and cross-module participation, but this effect was reversed in the cerebellar connections^38^. Hormones may also play a potential role in the sex discrepancies. Human and animal studies have found that reduced estrogen levels were associated with HD^39^, and our previous study has identified HD associations related to peripheral hormonally regulated tissues^10^. Beyond sex differences, most IDPs were positively genetically correlated with HD (i.e., larger volumes or more activation of specific brain regions linked to higher prevalence of HD), which suggested that hearing loss may lead to compensation of other activations or functions in the brain, such as increased demand of deciphering language through print and lip-reading, visual perception and processing, and spatial navigation^40,41^. However, we also observed other IDPs with negative HD genetic correlations, indicating brain atrophy or decreased activation of certain brain structures such as in somatosensory area BA2, the retrolenticular part of the internal capsule, and the temporal lobe. This may be related to the possibility that there may be shared aging-related neuropathologic processes that influence cochlear and central auditory function or that degraded auditory signals and alterations in auditory neural stimulation could lead to volume losses of other brain regions^42^. Further studies with respect to these genetic correlations are needed.

While previous studies used observational data to estimate the relationship of HD with changes in brain structure and function, we inferred possible causal relationships using LCV analysis with genetic information. In the sex-combined analysis, five IDPs had significant causal effects on HD: IDP T1 FAST ROIs R supramarg gyrus post, aparc-a2009s rh volume G-temp-sup-Plan-polar, rfMRI amplitudes ICA100 node 27, rfMRI connectivity ICA100 edge 1140, and aseg rh volume vessel. Interestingly, the supramarginal gyrus is involved in brain functions of receiving auditory inputs and phonological processing and the volume of grey matter in the right supramarginal gyrus was negatively associated with HD^43^. Similarly, a case-control study found that age-related hearing loss patients had deceased connectivity in the bilateral supramarginal gyrus^44^. In previous studies, individuals with HD showed a decreased volume in superior temporal gyrus, transverse temporal gyrus, and posterior superior temporal gyrus^14,15,45^. However, the present study found that the volume of planum polare in the anterior superior temporal gyrus had a positive relationship with HD. This may be due to the different functional focus of the superior temporal gyrus regions. As for two rfMRI IDPs, the fluctuation amplitudes of node 27 in dimensionality 100 were located in bilateral motor and premotor areas (Supplementary Fig. S2), while the partial correlation of edge 1140 in dimensionality 100 linked node 12 (right prefrontal area, right central parietal lobule, and left anterior and posterior lobes of cerebellum; Supplementary Fig. S3) and node 49 (bilateral inferior parietal lobule; Supplementary Fig. S4). The negative relationship of the two rfMRI traits with HD implied that auditory–motor integration and cerebellar–cerebral circuits may be involved in speech perception or compensate for impaired auditory processing^46,47^. In sex-specific analysis, the direction of most causal effects was from IDPs to HD, with a positive relationship (Fig. 2, Supplementary Tables S7, S8). However, in females, the volume of left hippocampus and the mean thickness of right caudal middle frontal gyrus showed a causal effect from HD to IDPs. And IDPs of left superior parietal lobule (white surface area), right cerebellum X (grey matter volume), and right middle temporal gyrus (white surface area) had a negative relationship with HD. In males, only the direction of the mean orientation dispersion index of left superior corona radiata was from HD to IDPs and four dMRI IDPs negatively related to HD. Unlike most previous studies that focused on the volumetric declines and functional changes of brain regions especially in auditory cortex due to HD, the present study used brain-wide information derived from multiple MRI modalities to expand our understanding of which brain-related mechanisms can be linked to auditory function decline^41,46^.

Among those causal associations found in the LCV analysis, seven were verified using the MR approach. These methods for genetically informed causal inference analysis are based on different assumptions and convergent results between them can be considered highly reliable. Vessel volume in the right hemisphere was the only IDP that demonstrated a negative causal effect on HD in the sex-combined MR analysis, meaning that the smaller the vessel volume, the higher the risk of developing HD. A previous case-control study showed that sudden sensorineural hearing loss was associated with vertebrobasilar insufficiency^48^, suggesting that changes in the circulatory system may have a potential role in auditory function, although sudden sensorineural hearing loss has a different pathological process than age-related hearing loss. The vertebrobasilar artery supplies blood to the brainstem, cerebellum, and vestibulocochlear organs, thus compromised vertebrobasilar arterial blood flow could lead to vestibulocochlear symptoms. In females, we found that the decreased left hippocampal volume was a causal factor of HD. This may be because the auditory–hippocampal interactions also contribute to perception and cognition^49,50^. Nevertheless, unlike the previous studies investigating the hippocampal alterations in patients with HD^51^, we revealed the causal effect of hippocampal volume changes on HD development. Another risk factor we identified was the reduced volume of grey matter in right X cerebellum (flocculonodular lobule), which is an essential node in the vestibular system. In addition to its role in motor function, the cerebellum is involved in vestibular and sensorimotor integration. However, whether the volume is positively or negatively correlated with HD remains unclear^41^. We also found that the increased volume of left V1 (primary visual cortex) white surface and the greater fluctuation amplitudes of node 46 in dimensionality 100 (bilateral middle temporal gyrus; Supplementary Fig. S5) had a positive relationship with HD. Both regions are involved in visual perception and processing^52^; notably, increased visual activation and connectivity were reported to be associated with auditory deprivation^46,53^. In males, we found that HD was associated with the increased mean thickness of right white surface and a reduced orientation dispersion index in the left superior corona radiata. Brain thickness has been sparsely studied compared to grey matter and white matter volume^41^. However, a few previous studies showed increased cortical thickness in the right precuneus and the left posterior cingulate gyrus in deaf patients^41^, which may be due to the compensatory plasticity to auditory impairment^54^. Our MR result on orientation dispersion index in left superior corona radiata was consistent with the findings of the LCV analysis that fractional anisotropy, diffusion tensor mode, and L1 have a positive correlation with HD. However, two IDPs had opposite causal directions in the MR and LCV analyses: the volume of left hippocampus and the fluctuation amplitudes of node 46, possibly suggesting a bidirectional causal relationship with HD. Importantly, the signs of the causal effects estimated using multivariable regression were consistent with the signs in MR analysis, strengthening the finding of a causal relationships between IDPs and HD.

Our local genetic correlation analysis provided evidence supporting the shared genetic hypothesis for brain structure and function and HD. In all, we discovered 13 semi-independent loci having pleiotropic associations between the seven IDPs and HD, and three of them showed significant local genetic correlation at more than one locus. Interestingly, several local genetic correlations showed opposite signs of genetic correlation at different loci for a specific IDP. For example, the volume of vessel in the right hemisphere had a strongly positive correlation at locus 965 (local r_g_ = 0.942) and locus 966 (local r_g_ = 0.881), but a negative correlation at locus 1280 (local r_g_ = −0.950). This suggests that their pleiotropic relationship in different brain regions may have different genetic mechanisms. In colocalization analysis, two SNP signals were recognized, although the PPFC was not more than 0.7. We found that rs5899177 was the causal SNP shared by the volume of left hippocampus and HD in females. This SNP is an eQTL in multiple tissues. However, we did not find associated transcriptomic regulation that shared the causal SNP with the IDP and HD in this study. We also observed a colocalization signal for the rs13026575 shared by the volume of left primary visual cortex, HD, and *SPTBN1* transcriptomic regulation in brain tissues in females. *SPTBN1* encodes βII-spectrin that forms cell membrane cytoskeleton, and the deficiency of neuronal βII-spectrin could cause measurable compromise of neural development and function^55^. This gene was associated with age-related hearing loss in a previous GWAS meta-analysis^56^, providing a potential explanation for the different IDP–HD associations between sexes^10^.

Leveraging GWAS data from different cohorts, the present study took multiple steps to systematically investigate HD pleiotropy with brain structure and function and identified causal relationships between them for the first time. However, this study has several limitations. First, the brain regions analyzed were not necessarily anatomically defined to address language processing, such as the cortical areas specialized in the auditory pathway, written language, and lip-reading. The only structural IDP of temporal lobe identified with causal effect was the right area of middle temporal gyrus in female LCV analysis. To improve the statistical power, future studies could benefit from repeat imaging scans with larger sample sizes that will be released from the UKB. Moreover, we cannot distinguish sub-types of hearing loss based on the available data. However, age-related HD is the most common type and accounts for the majority of HD^57^. Thus, the IDPs identified in the present study are likely connected to age-related HD. In addition, due to the cross-sectional study design in the multivariable generalized linear regression analysis because of the relatively small sample size in the follow-up imaging visit in the UKB, causal inferences for the IDP–HD associations cannot be made. Although the effects estimated in multivariable generalized linear regression analysis were consistent with those in MR, the IDPs identified to have a causal relationship with HD in the current study should be further validated using large-scale longitudinal cohort studies with brain imaging. Finally, due to the different approaches we used, the causal SNPs detected with HyPrColoc analysis did not locate within the loci identified with LAVA analysis. LAVA aimed to find genetic regions where IDPs and HD correlated, while HyPrColoc was used to find single shared SNP in a region. Therefore, the region containing variants with small pleiotropy effects may not have a causal SNP that reaches the criteria of the HyPrColoc. Despite PPFC estimates for loci in HyPrColoc being less than 0.7, two regions showed significant colocalization signals according to this method.

In conclusion, this study reports comprehensive evidence of genetic correlations and causal associations of brain structure and function with HD, and highlights sex-specific pleiotropic mechanisms shared between them. The findings may facilitate the identification of diagnostic and therapeutic targets in HD population, considering differences between females and males.

## Data Availability

All data produced in the present work are contained in the manuscript.

## ABBREVIATIONS

dMRI: Diffusion MRI
eQTL: Expression quantitative trait loci
FDR: False discovery rate
gcp: Genetic causality proportions
HD: Hearing difficulty
HyPrColoc: Hypothesis prioritization in multi-trait colocalization
IDPs: Imaging-derived phenotypes
IVs: Instrumental variables
IVW: Inverse variance weighting
LAVA: Local analysis of [co]variant association
LCV: Latent causal variable
LD: Linkage disequilibrium
LDSC: Linkage disequilibrium score regression
MAF: Minor allele frequency
MRI: Magnetic resonance imaging
PPFC: Posterior probability of full colocalization
P_R_: Regional association probability
P_snp_: PPFC explained by the candidate causal variant
r_g_: Genetic correlations
rfMRI: Resting-state functional MRI
SWI: Susceptibility-weighted imaging
tfMRI: Task functional MRI
YLDs: Years lived with disability

## ACKNOWLEDGEMENTS

We are grateful to the participants and investigators of the UKB, NHS I, NHS II, and HPFS studies.

## AUTHORS’ CONTRIBUTIONS

JH and RP designed the study. JH conducted the primary data analysis. BCM, FDA, GAP, DK, SGC, GCC, APM, and CHvD contributed to the data interpretation. All the authors participated in the interpretation of data and critical revision of the manuscript for important intellectual content. RP obtained the primary funding. RP supervised the study. All authors read and approved the final manuscript.

## DATA AND CODE AVAILABILITY

The UKB individual data could be obtained by applying on the website (http://www.ukbiobank.ac.uk/). GWAS data were free available (Hearing difficulty: https://doi.org/10.5281/zenodo.7897038; Brain imaging-derived phenotypes: https://open.win.ox.ac.uk/ukbiobank/big40/). eQTL data were extracted from the GTEx v.8 release (https://gtexportal.org/home/). Global genetic correlations were estimated using LDSC v1.0.1 (https://github.com/bulik/ldsc). Genetic causality proportions were calculated using LCV (https://github.com/lukejoconnor/LCV). MR analyses were performed using MRlap v0.0.3 (https://github.com/n-mounier/MRlap). Local genetic correlations were computed using LAVA v0.1.0 (https://github.com/josefin-werme/LAVA). Colocalization analyses were performed using HyPrColoc v1.0 (https://github.com/jrs95/hyprcoloc).

## SUPPLEMENTARY FILES

Supplementary Fig. S1: Sex differences in the genetic correlation between hearing difficulty (HD) and brain imaging-derived phenotypes (IDPs). The blue line corresponds to the linear fit (r_Pearson_ = 0.21, *P* < 2.2 × 10^-16^) of the relationship between females and males. Labels are reported for the three IDPs with the significant sex difference in HD genetic correlation after Bonferroni correction (*P* < 0.05/3024). Full results are available in Supplementary Table S5.

Supplementary Fig. S2: Node 27 of dimensionality 100 separated by spatial ICA in resting-state functional magnetic resonance imaging.

Supplementary Fig. S3: Node 12 of dimensionality 100 separated by spatial ICA in resting-state functional magnetic resonance imaging.

Supplementary Fig. S4: Node 49 of dimensionality 100 separated by spatial ICA in resting-state functional magnetic resonance imaging.

Supplementary Fig. S5: Node 46 of dimensionality 100 separated by spatial ICA in resting-state functional magnetic resonance imaging.

Supplementary Table S1: Information for brain imaging-derived phenotypes (IDPs).

Supplementary Table S2: Global genetic correlation between hearing difficulty (HD) and brain imaging-derived phenotypes (IDPs) in the sex-combined analysis.

Supplementary Table S3: Global genetic correlation between hearing difficulty (HD) and brain imaging-derived phenotypes (IDPs) in females.

Supplementary Table S4: Global genetic correlation between hearing difficulty (HD) and brain imaging-derived phenotypes (IDPs) in males.

Supplementary Table S5: Sex differences in the genetic correlation between hearing difficulty (HD) and brain imaging-derived phenotypes (IDPs).

Supplementary Table S6: Putative causal effects between hearing difficulty (HD) and brain imaging-derived phenotypes (IDPs) in the sex-combined latent causal variable analysis.

Supplementary Table S7: Putative causal effects between hearing difficulty (HD) and brain imaging-derived phenotypes (IDPs) in females using latent causal variable analysis.

Supplementary Table S8: Putative causal effects between hearing difficulty (HD) and brain imaging-derived phenotypes (IDPs) in males using latent causal variable analysis.

Supplementary Table S9: Results of sex-combined Mendelian randomization analysis for hearing difficulty (HD) and brain imaging-derived phenotypes (IDPs).

Supplementary Table S10: Results of Mendelian randomization analysis for hearing difficulty (HD) and brain imaging-derived phenotypes (IDPs) in females.

Supplementary Table S11: Results of Mendelian randomization analysis for hearing difficulty (HD) and brain imaging-derived phenotypes (IDPs)in males.

Supplementary Table S12: Results of multivariable generalized linear regression analysis for hearing difficulty (HD) and brain imaging-derived phenotypes (IDPs) identified in the Mendelian randomization analysis.

Supplementary Table S13: Local genetic correlation between hearing difficulty (HD) and brain imaging-derived phenotypes (IDPs) identified in the Mendelian randomization analysis.

Supplementary Table S14: Colocalization between hearing difficulty (HD) and brain imaging-derived phenotypes (IDPs) identified in the female Mendelian randomization analysis.

Supplementary Table S15: Expression quantitative trait loci data used in colocalization analysis.

Supplementary Table S16: Colocalization between hearing difficulty (HD), brain imaging-derived phenotype of volume of left hippocampus (IDP 0199: aseg lh volume Hippocampus), and *NINJ1* transcriptomic regulation in brain tissues from GTEx v.8 release.

Supplementary Table S17: Colocalization between hearing difficulty (HD), brain imaging-derived phenotype of volume of left primary visual cortex (IDP 0419: BA-exvivo lh volume V1), and three genes transcriptomic regulation in brain tissues from GTEx v.8 release.

## REFERENCES

1. Chadha S, Kamenov K, Cieza A. The world report on hearing, 2021. Bull World Health Organ. 2021;99(4):242–242A. doi:10.2471/BLT.21.285643

2. GBD 2019 Hearing Loss Collaborators. Hearing loss prevalence and years lived with disability, 1990-2019: findings from the Global Burden of Disease Study 2019. LANCET. 2021;397(10278):996–1009. doi:10.1016/S0140-6736(21)00516-X

3. McDaid D, Park AL, Chadha S. Estimating the global costs of hearing loss. Int J Audiol. 2021;60(3):162–170. doi:10.1080/14992027.2021.1883197

4. GBD 2019 Ageing Collaborators. Global, regional, and national burden of diseases and injuries for adults 70 years and older: systematic analysis for the Global Burden of Disease 2019 Study. BMJ. 2022;376:e068208. doi:10.1136/bmj-2021-068208

5. Olusanya BO, Neumann KJ, Saunders JE. The global burden of disabling hearing impairment: a call to action. Bull World Health Organ. 2014;92(5):367–73. doi:10.2471/BLT.13.128728

6. Nordvik O, Laugen Heggdal PO, Brannstrom J, Vassbotn F, Aarstad AK, Aarstad HJ. Generic quality of life in persons with hearing loss: a systematic literature review. BMC Ear Nose Throat Disord. 2018;18:1. doi:10.1186/s12901-018-0051-6

7. Jiang F, Mishra SR, Shrestha N, et al. Association between hearing aid use and all-cause and cause-specific dementia: an analysis of the UK Biobank cohort. Lancet Public Health. 2023;8(5):e329–e338. doi:10.1016/S2468-2667(23)00048-8

8. Huang HM, Chen GS, Liu ZY, et al. Age-related hearing loss accelerates the decline in fast speech comprehension and the decompensation of cortical network connections. Neural Regen Res. 2023;18(9):1968–1975. doi:10.4103/1673-5374.361530

9. Xu S, Hou C, Han X, et al. Adverse health consequences of undiagnosed hearing loss at middle age: A prospective cohort study with the UK Biobank. MATURITAS. 2023;174:30–38. doi:10.1016/j.maturitas.2023.05.002

10. De Angelis F, Zeleznik OA, Wendt FR, et al. Sex differences in the polygenic architecture of hearing problems in adults. Genome Med. 2023;15(1):36. doi:10.1186/s13073-023-01186-3

11. World Health Organization. World report on hearing. 2021. https://apps.who.int/iris/handle/10665/339913

12. Boyen K, Langers DR, de Kleine E, van Dijk P. Gray matter in the brain: differences associated with tinnitus and hearing loss. Hear Res. 2013;295:67–78. doi:10.1016/j.heares.2012.02.010

13. Rosemann S, Thiel CM. Neuroanatomical changes associated with age-related hearing loss and listening effort. Brain Struct Funct. 2020;225(9):2689–2700. doi:10.1007/s00429-020-02148-w

14. Shibata DK. Differences in brain structure in deaf persons on MR imaging studied with voxel-based morphometry. AJNR Am J Neuroradiol. 2007;28(2):243–9.

15. Simon M, Campbell E, Genest F, MacLean MW, Champoux F, Lepore F. The Impact of Early Deafness on Brain Plasticity: A Systematic Review of the White and Gray Matter Changes. Front Neurosci. 2020;14:206. doi:10.3389/fnins.2020.00206

16. Yang M, Chen HJ, Liu B, et al. Brain structural and functional alterations in patients with unilateral hearing loss. Hear Res. 2014;316:37–43. doi:10.1016/j.heares.2014.07.006

17. Golub JS. Brain changes associated with age-related hearing loss. Curr Opin Otolaryngol Head Neck Surg. 2017;25(5):347–352. doi:10.1097/MOO.0000000000000387

18. Li Y, Ding G, Booth JR, et al. Sensitive period for white-matter connectivity of superior temporal cortex in deaf people. Hum Brain Mapp. 2012;33(2):349–59. doi:10.1002/hbm.21215

19. Propst EJ, Greinwald JH, Schmithorst V. Neuroanatomic differences in children with unilateral sensorineural hearing loss detected using functional magnetic resonance imaging. Arch Otolaryngol Head Neck Surg. 2010;136(1):22–6. doi:10.1001/archoto.2009.208

20. Bycroft C, Freeman C, Petkova D, et al. The UK Biobank resource with deep phenotyping and genomic data. NATURE. 2018;562(7726):203–209. doi:10.1038/s41586-018-0579-z

21. Bao Y, Bertoia ML, Lenart EB, et al. Origin, Methods, and Evolution of the Three Nurses’ Health Studies. Am J Public Health. 2016;106(9):1573–81. doi:10.2105/AJPH.2016.303338

22. Shargorodsky J, Curhan SG, Eavey R, Curhan GC. A prospective study of cardiovascular risk factors and incident hearing loss in men. LARYNGOSCOPE. 2010;120(9):1887–91. doi:10.1002/lary.21039

23. Smith SM, Douaud G, Chen W, et al. An expanded set of genome-wide association studies of brain imaging phenotypes in UK Biobank. Nat Neurosci. 2021;24(5):737–745. doi:10.1038/s41593-021-00826-4

24. Bulik-Sullivan BK, Loh PR, Finucane HK, et al. LD Score regression distinguishes confounding from polygenicity in genome-wide association studies. Nat Genet. 2015;47(3):291–5. doi:10.1038/ng.3211

25. Bulik-Sullivan B, Finucane HK, Anttila V, et al. An atlas of genetic correlations across human diseases and traits. Nat Genet. 2015;47(11):1236–41. doi:10.1038/ng.3406

26. Genomes Project C, Auton A, Brooks LD, et al. A global reference for human genetic variation. NATURE. 2015;526(7571):68–74. doi:10.1038/nature15393

27. International HapMap C. The International HapMap Project. NATURE. 2003;426(6968):789–96. doi:10.1038/nature02168

28. O’Connor LJ, Price AL. Distinguishing genetic correlation from causation across 52 diseases and complex traits. Nat Genet. 2018;50(12):1728–1734. doi:10.1038/s41588-018-0255-0

29. Mounier N, Kutalik Z. Bias correction for inverse variance weighting Mendelian randomization. Genet Epidemiol. 2023;47(4):314–331. doi:10.1002/gepi.22522

30. Hemani G, Zheng J, Elsworth B, et al. The MR-Base platform supports systematic causal inference across the human phenome. eLife. 2018;7doi:10.7554/eLife.34408

31. Sanderson E, Glymour MM, Holmes MV, et al. Mendelian randomization. Nature Reviews Methods Primers. 2022;2(1)doi:10.1038/s43586-021-00092-5

32. Hemani G, Tilling K, Davey Smith G. Orienting the causal relationship between imprecisely measured traits using GWAS summary data. PLoS Genet. 2017;13(11):e1007081. doi:10.1371/journal.pgen.1007081

33. Sterne JA, Davey Smith G. Sifting the evidence-what’s wrong with significance tests? BMJ. 2001;322(7280):226–31. doi:10.1136/bmj.322.7280.226

34. Werme J, van der Sluis S, Posthuma D, de Leeuw CA. An integrated framework for local genetic correlation analysis. Nat Genet. 2022;54(3):274–282. doi:10.1038/s41588-022-01017-y

35. Foley CN, Staley JR, Breen PG, et al. A fast and efficient colocalization algorithm for identifying shared genetic risk factors across multiple traits. Nat Commun. 2021;12(1):764. doi:10.1038/s41467-020-20885-8

36. Consortium GT. The GTEx Consortium atlas of genetic regulatory effects across human tissues. SCIENCE. 2020;369(6509):1318–1330. doi:10.1126/science.aaz1776

37. Han B, Eskin E. Interpreting meta-analyses of genome-wide association studies. PLoS Genet. 2012;8(3):e1002555. doi:10.1371/journal.pgen.1002555

38. Ingalhalikar M, Smith A, Parker D, et al. Sex differences in the structural connectome of the human brain. Proc Natl Acad Sci U S A. 2014;111(2):823–8. doi:10.1073/pnas.1316909110

39. Curhan SG, Eliassen AH, Eavey RD, Wang M, Lin BM, Curhan GC. Menopause and postmenopausal hormone therapy and risk of hearing loss. Menopause. 2017;24(9):1049–1056. doi:10.1097/GME.0000000000000878

40. Alfandari D, Vriend C, Heslenfeld DJ, Versfeld NJ, Kramer SE, Zekveld AA. Brain Volume Differences Associated With Hearing Impairment in Adults. Trends Hear. 2018;22:2331216518763689. doi:10.1177/2331216518763689

41. Gregoire A, Deggouj N, Dricot L, Decat M, Kupers R. Brain Morphological Modifications in Congenital and Acquired Auditory Deprivation: A Systematic Review and Coordinate-Based Meta-Analysis. Front Neurosci. 2022;16:850245. doi:10.3389/fnins.2022.850245

42. Lin FR, Ferrucci L, An Y, et al. Association of hearing impairment with brain volume changes in older adults. NEUROIMAGE. 2014;90:84–92. doi:10.1016/j.neuroimage.2013.12.059

43. Hartwigsen G, Baumgaertner A, Price CJ, Koehnke M, Ulmer S, Siebner HR. Phonological decisions require both the left and right supramarginal gyri. Proc Natl Acad Sci U S A. 2010;107(38):16494–9. doi:10.1073/pnas.1008121107

44. Zhu S, Song J, Xia W, Xue Y. Aberrant brain functional network strength related to cognitive impairment in age-related hearing loss. Front Neurol. 2022;13:1071237. doi:10.3389/fneur.2022.1071237

45. Manno FAM, Rodriguez-Cruces R, Kumar R, Ratnanather JT, Lau C. Hearing loss impacts gray and white matter across the lifespan: Systematic review, meta-analysis and meta-regression. NEUROIMAGE. 2021;231:117826. doi:10.1016/j.neuroimage.2021.117826

46. Slade K, Plack CJ, Nuttall HE. The Effects of Age-Related Hearing Loss on the Brain and Cognitive Function. Trends Neurosci. 2020;43(10):810–821. doi:10.1016/j.tins.2020.07.005

47. Xu XM, Jiao Y, Tang TY, et al. Dissociation between Cerebellar and Cerebral Neural Activities in Humans with Long-Term Bilateral Sensorineural Hearing Loss. Neural Plast. 2019;2019:8354849. doi:10.1155/2019/8354849

48. Hsu YH, Hu HY, Chiu YC, Lee FP, Huang HM. Association of Sudden Sensorineural Hearing Loss With Vertebrobasilar Insufficiency. JAMA Otolaryngol Head Neck Surg. 2016;142(7):672–5. doi:10.1001/jamaoto.2016.0845

49. Armstrong NM, An Y, Doshi J, et al. Association of Midlife Hearing Impairment With Late-Life Temporal Lobe Volume Loss. JAMA Otolaryngol Head Neck Surg. 2019;145(9):794–802. doi:10.1001/jamaoto.2019.1610

50. Billig AJ, Lad M, Sedley W, Griffiths TD. The hearing hippocampus. Prog Neurobiol. 2022;218:102326. doi:10.1016/j.pneurobio.2022.102326

51. Zhang L, Wang J, Sun H, Feng G, Gao Z. Interactions between the hippocampus and the auditory pathway. Neurobiol Learn Mem. 2022;189:107589. doi:10.1016/j.nlm.2022.107589

52. Onitsuka T, Shenton ME, Salisbury DF, et al. Middle and inferior temporal gyrus gray matter volume abnormalities in chronic schizophrenia: an MRI study. Am J Psychiatry. 2004;161(9):1603–11. doi:10.1176/appi.ajp.161.9.1603

53. Yang T, Liu Q, Fan X, Hou B, Wang J, Chen X. Altered regional activity and connectivity of functional brain networks in congenital unilateral conductive hearing loss. Neuroimage Clin. 2021;32:102819. doi:10.1016/j.nicl.2021.102819

54. Alzaher M, Vannson N, Deguine O, Marx M, Barone P, Strelnikov K. Brain plasticity and hearing disorders. Rev Neurol (Paris). 2021;177(9):1121–1132. doi:10.1016/j.neurol.2021.09.004

55. Cousin MA, Creighton BA, Breau KA, et al. Pathogenic SPTBN1 variants cause an autosomal dominant neurodevelopmental syndrome. Nat Genet. 2021;53(7):1006–1021. doi:10.1038/s41588-021-00886-z

56. Nagtegaal AP, Broer L, Zilhao NR, et al. Genome-wide association meta-analysis identifies five novel loci for age-related hearing impairment. Sci Rep. 2019;9(1):15192. doi:10.1038/s41598-019-51630-x

57. Zahnert T. The differential diagnosis of hearing loss. Dtsch Arztebl Int. 2011;108(25):433–43; quiz 444. doi:10.3238/arztebl.2011.0433

