## Supplementary Figures for "Sex differences in the pleiotropy of hearing difficulty with imaging-derived phenotypes: a brain-wide investigation"

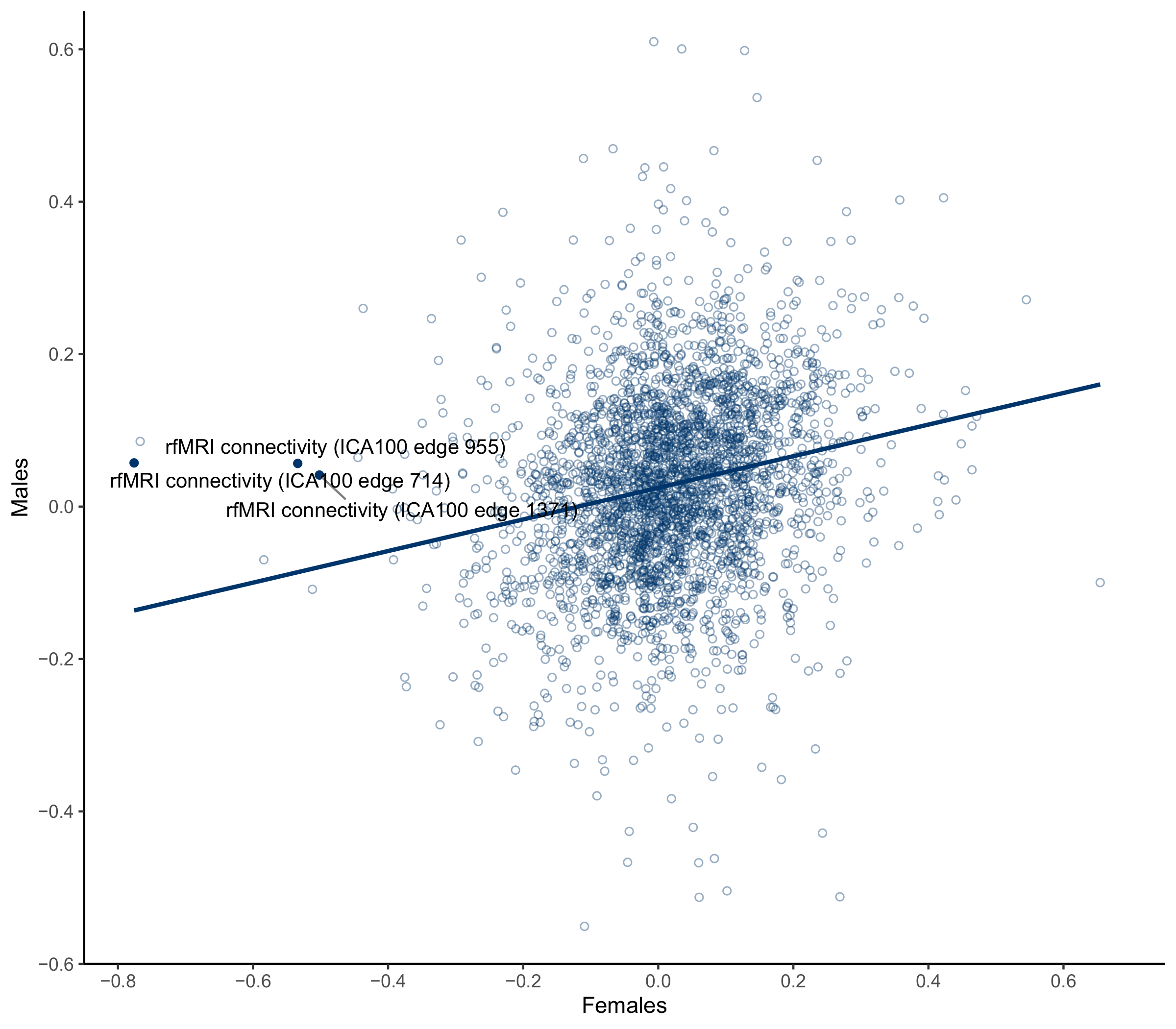

Supplementary Fig. S1: Sex differences in the genetic correlation between hearing difficulty (HD) and brain imaging-derived phenotypes (IDPs). The blue line corresponds to the linear fit (r_Pearson_ = 0.21, *P* < 2.2 × 10^-16^) of the relationship between females and males. Labels are reported for the three IDPs with the significant sex difference in HD genetic correlation after Bonferroni correction (*P* < 0.05/3024). Full results are available in Supplementary Table S5.

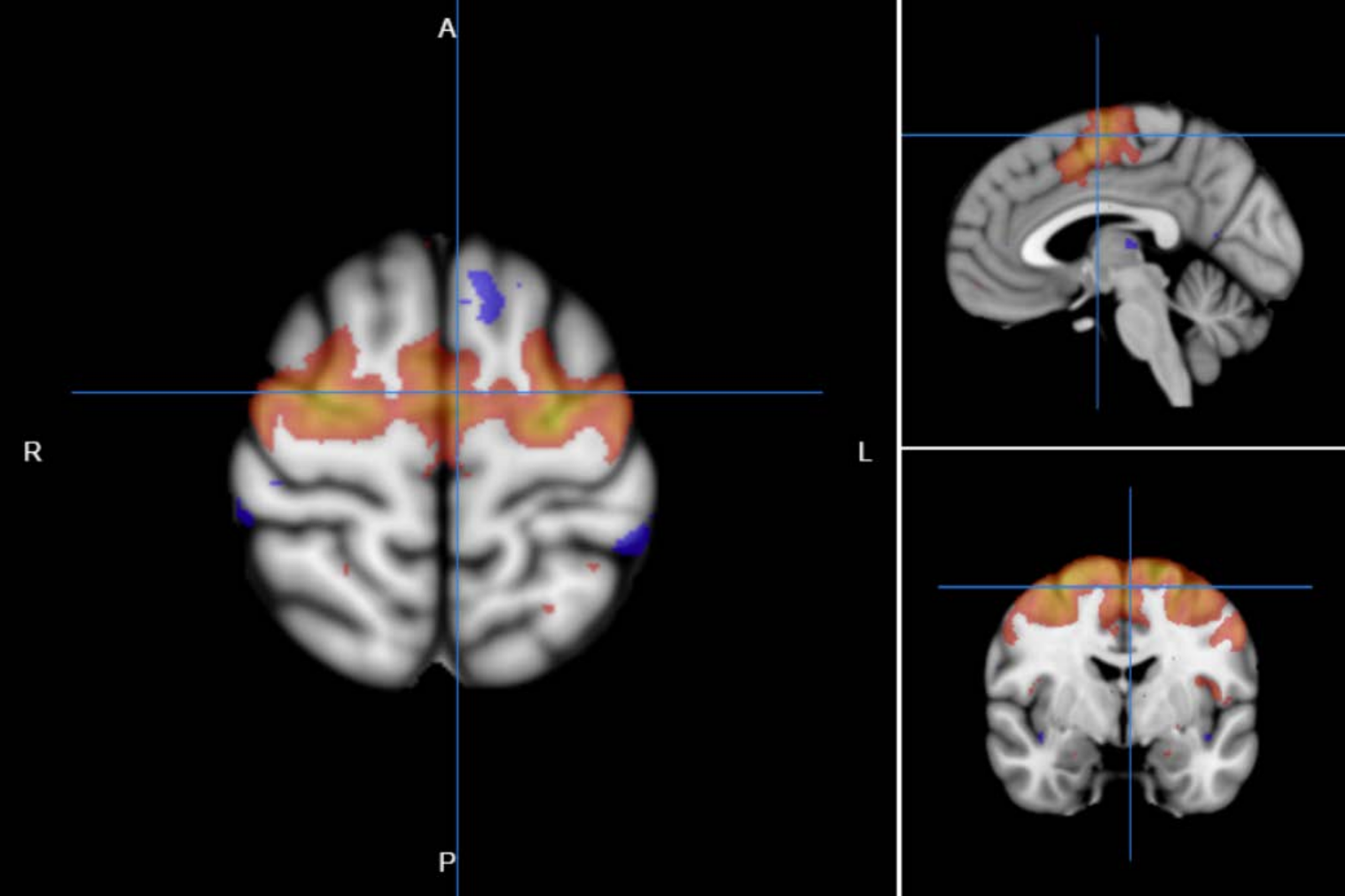

Supplementary Fig. S2: Node 27 of dimensionality 100 separated by spatial ICA in resting-state functional magnetic resonance imaging.

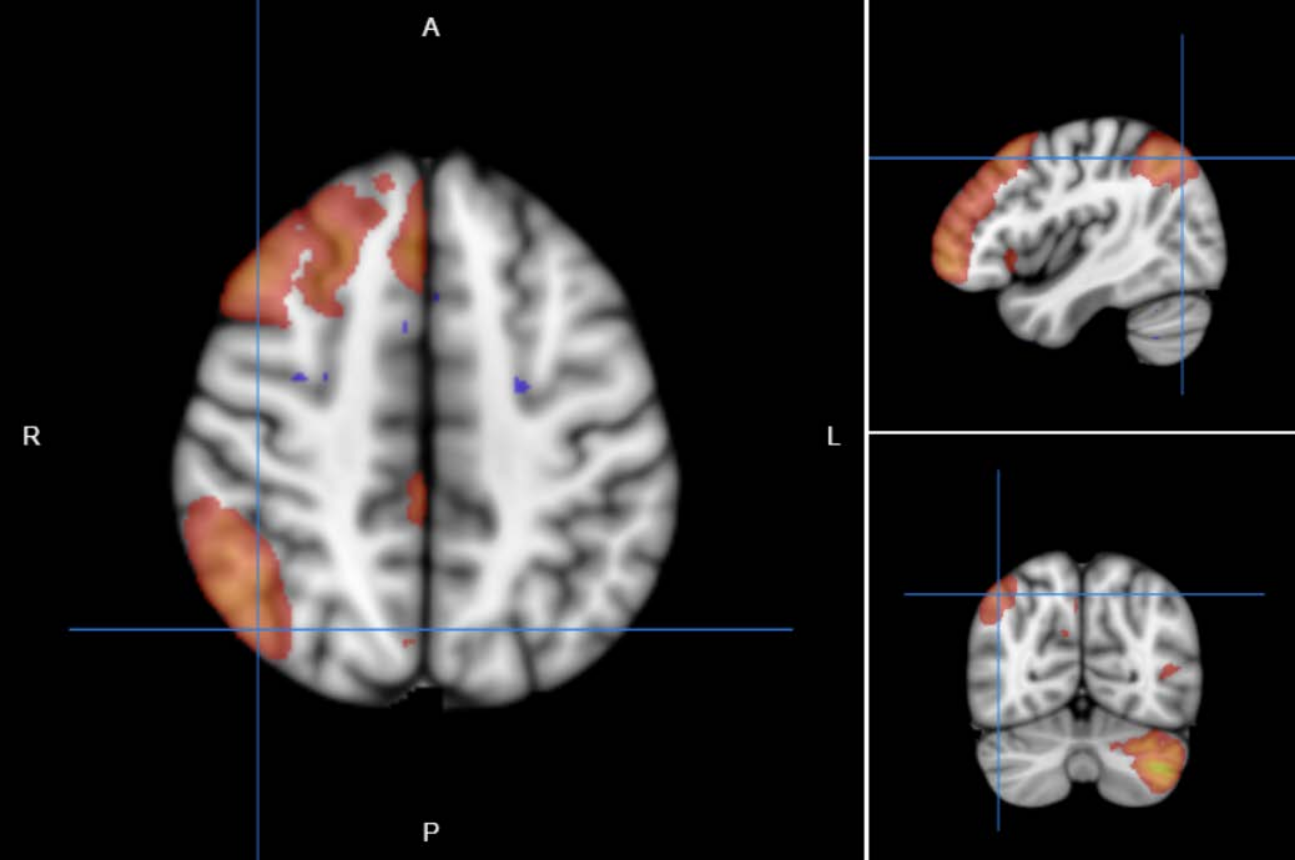

Supplementary Fig. S3: Node 12 of dimensionality 100 separated by spatial ICA in resting-state functional magnetic resonance imaging.

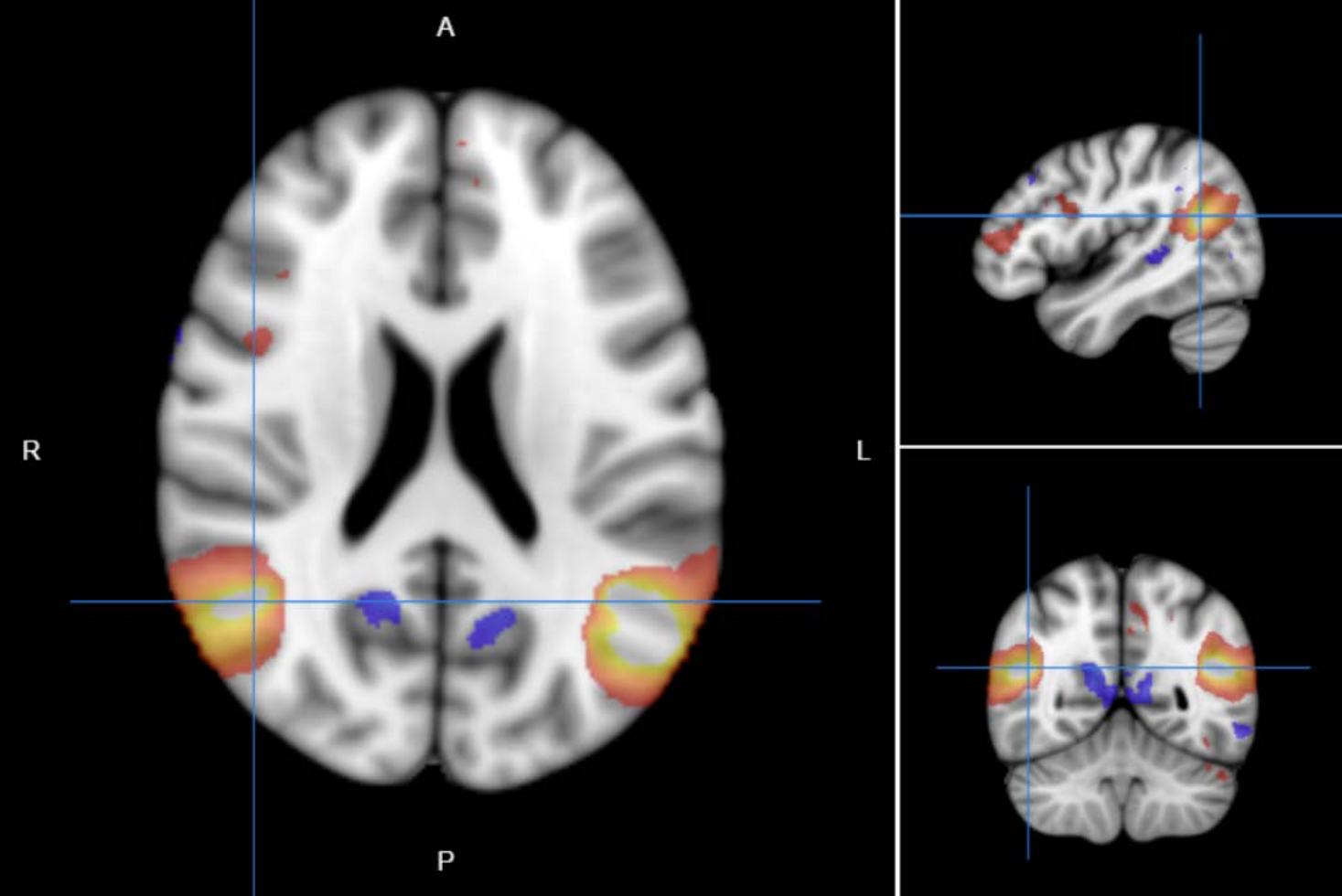

Supplementary Fig. S4: Node 49 of dimensionality 100 separated by spatial ICA in resting-state functional magnetic resonance imaging.

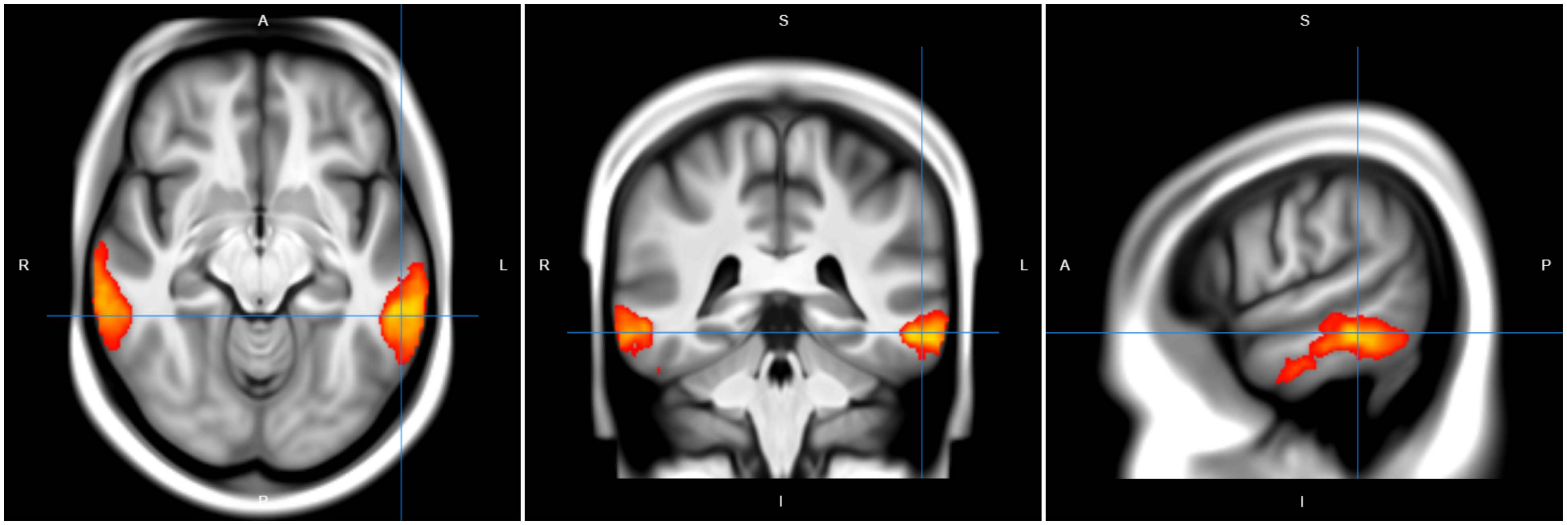

Supplementary Fig. S5: Node 46 of dimensionality 100 separated by spatial ICA in resting-state functional magnetic resonance imaging.
